# Time to full enteral feeds in hospitalised preterm and very low birth weight infants in Nigeria and Kenya

**DOI:** 10.1101/2022.11.04.22281964

**Authors:** Zainab O Imam, Helen M Nabwera, Olukemi O Tongo, Pauline EA Andang’o, Isa Abdulkadir, Chinyere V Ezeaka, Beatrice N Ezenwa, Iretiola B Fajolu, Martha K Mwangome, Dominic D Umoru, Abimbola E Akindolire, Walter Otieno, Macrine Olwala, Grace M Nalwa, Alison W Talbert, Ismaela Abubakar, Nicholas D Embleton, Stephen J Allen, the Neonatal Nutrition Network (NeoNuNet)

## Abstract

**Background:** Preterm (born < 37 weeks’ gestation) and very low birthweight (VLBW; <1.5kg) infants are at the greatest risk of morbidity and mortality within the first 28 days of life. Establishing full enteral feeds is a vital aspect of their clinical care. Evidence predominantly from high income countries shows that early and rapid advancement of feeds is safe and reduces length of hospital stay and adverse health outcomes. However, there are limited data on feeding practices and factors that influence the advancement of feeds among these vulnerable infants in sub-Saharan Africa.

**Aim:** To identify factors that influence the time to full enteral feeds, defined as tolerance of 120ml/kg/day, in hospitalised preterm and VLBW infants in neonatal units in two sub-Saharan African countries.

**Methods:** Demographic and clinical variables were collected for newborns admitted to 7 neonatal units in Nigeria and Kenya over 6-months. Multiple linear regression analysis was conducted to identify factors independently associated with time to full enteral feeds.

**Results:** Of the 2280 newborn infants admitted, 484 were preterm and VLBW. Overall, 222/484 (45.8%) infants died with over half of the deaths (136/222; 61.7%) occurring before the first feed. The median (inter-quartile range) time to first feed was 46 (27, 72) hours of life and time to full enteral feeds (tFEF) was 8 (4.5,12) days with marked variation between neonatal units. Independent predictors of tFEF were time to first feed (unstandardised coefficient B 1.75; 95% CI 1.16 to 2.34; p value <0.001) and the occurrence of respiratory distress (−1.89; −3.27 to −0.5; <0.007) and necrotising enterocolitis (4.59; 1.16 to 7.92; <0.009).

**Conclusion:** The use of standardised feeding guidelines may decrease variations in clinical practice, shorten tFEF and thereby improve newborn outcomes.

## Introduction

Complications of prematurity (birth before 37 completed weeks) are the leading cause of neonatal and under 5 mortality (1), with the frequency and severity of morbidity and mortality increasing with decreasing gestational age (GA) and birthweight (2). In a recent review of hospitalised newborns in Nigeria and Kenya, the leading risk factors for mortality were extreme prematurity (<28 weeks’ gestation) and very low birthweight (VLBW; <1.5kg) which increased the odds of mortality by 12 and 7 times respectively (3).

Early enteral feeding with breastmilk is vital in reducing adverse outcomes among hospitalised preterm VLBW infants. In addition to providing essential macro- and micronutrients, breast milk promotes the maturation of the gut microbiome which, in turn, promotes immune modulation, digestion and metabolism of feeds and neurodevelopment (4). Early nutrition is therefore central to the prevention of short-term morbidities such as late-onset sepsis and feed intolerance with no evidence of an increase in necrotising enterocolitis (NEC); as well as promoting early infant growth, survival and long-term optimal neurodevelopmental outcomes (5, 6).

Enteral feeding and prematurity are the major risk factors for (NEC), a severe illness characterised by inflammation of the preterm infant gut leading to necrosis and/or perforation with a case fatality rate of 20-30% (7). To prevent NEC, many clinicians delay the commencement of enteral feeds and/or advance feed volumes slowly for very preterm/VLBW infants, although there are few data to show this reduces the risk of NEC (8). Despite the strict feeding protocols prescribed by Brown and Sweet (9), infection control measures and other practice changes made in neonatal intensive care, up to 7 - 10% of preterm infants in the United States and Canada continue to develop NEC (7, 8, 10) which has encouraged further research into optimal feeding regimens.

In very preterm infants (<32 weeks’ gestation), the inability to coordinate suck, swallow and breathing and high risk of feeding intolerance and NEC make early feeding challenging (5). In addition, breastmilk availability may be limited because of delayed lactogenesis due to maternal illness or a lack of adequate lactation support (11). Early feeding for preterm infants may involve cup, spoon or naso- or orogastric tube feeding with graduated volumes of expressed mother’s own or donor breastmilk or artificial milk formula (5, 12). In some neonatal units (NNUs) in low-and-middle-income countries (LMICs), donor human milk may be obtained from a healthy wet-nurse (13), or from human-milk banks although this is more prevalent in high-income countries; HICs) (14). Parenteral nutrition is often not available in LMICs, although intravenous fluids may still be used to provide energy and water until sufficient milk feeds are established (5).

The WHO infant feeding guideline group conducted systematic reviews comparing early versus delayed enteral feeding, and slow versus fast rates of feed advancement among preterm/VLBW infants, from which guidelines for optimal feeding of LBW infants in LMICs were developed (12). They recommend that “VLBW infants in LMICs be given 10ml/kg/day of enteral feeds preferably starting from the first day of life, with the remaining fluid requirement met by intravenous fluids. Feed volumes can be increased by up to 30ml/kg/day with careful monitoring for feed intolerance.” A subsequent Cochrane systematic review (15) with bi-annual overviews of the systematic review (16-19) comparing the effect of slow (15-24ml/kg/day) versus fast (30-40ml/kg/day) enteral feed advancement have confirmed that rapid advancement of feeds does not increase the incidence of NEC or mortality among preterm VLBW infants. Despite this evidence-base, management of feeds among hospitalised preterm/VLBW infants varies within and outside sub-Saharan Africa, which may contribute to poor outcomes (11).

The time to full enteral feeds (tFEF) can be defined as the time it takes for the infant to tolerate an adequate volume of enteral feeds such that intravenous fluids or parenteral nutrition can be discontinued. It is a quantifiable and objective measure of feeding practice that captures the complex interactions between several factors including the baseline clinical status of the infant, the feeding guidelines being followed, and the morbidities the infant may develop in the course of their admission. It is a sum of the effects of these factors on feed tolerance, encompassing what is prescribed versus what is actually tolerated by an infant. This study aimed to identify factors associated with the attainment of full enteral feeds in hospitalised preterm/VLBW infants across 7 neonatal units (NNUs) in Nigeria and Kenya.

## Methods

### Study design

Enteral feeding data of preterm, VLBW infants were collected prospectively in a multi-centre observational study (Neonatal Nutrition Network project, NeoNuNet; https://www.lstmed.ac.uk/nnu).

### Study setting

Details of the study setting have been described previously (3). The 7 NNUs consist of two secondary and five tertiary level referral units, serving predominantly urban populations. They offer newborn care services to both inborn (born within participating perinatal centres) and outborn (born elsewhere then referred in) infants and provide newborn resuscitation, oxygen therapy, modified bubble continuous positive airway pressure (bCPAP), intravenous fluids, preterm enteral feeding, incubator care, intermittent and/or continuous Kangaroo Care and phototherapy; none of the units routinely used CPAP machines or mechanical ventilators. The NNUs were selected based on existing research and clinical training collaborative partnerships between the Liverpool School of Tropical Medicine (LSTM) co-investigators and NNU leads in Nigeria and Kenya.

### Study population and sampling

Data on all newborn infants, aged <48hours, admitted into the NNUs over any 6-month period between September 2018 and April 2019 were collected. There were no exclusion criteria; patients with congenital anomalies and those who were products of multiple gestation were included. Routine clinical information was collected until discharge, transfer or death.

### Data collection and management

Standardised case record forms, co-developed by NeoNuNet co-investigators based on national and WHO neonatal clinical guidelines (3), were piloted at all sites and refined prior to data collection. Data recorded included maternal pregnancy/birth and neonatal characteristics, feeding practices and morbidity and mortality. Common morbidities (asphyxia, respiratory distress, sepsis, feed intolerance and NEC) were diagnosed based on clinical features and results of laboratory and radiological investigations (where available).

Each NNU entered anonymised data into a REDCap (Research Electronic Data Capture) database, built and managed by a data manager at LSTM. All the data were exported into a .csv file to apply the INTERGROWTH standards (details below) to determine the appropriateness of growth for GA and sex. The database was imported into IBM Statistical Packages for Social Sciences software version 25 (SPSSv25) for analysis.

### Statistical analysis

VLBW was defined as birthweight between 1.0 and 1.499kg and extreme low birthweight (ELBW) as birthweight < 1.0 kg; the term VLBW refers to both VLBW and ELBW unless otherwise stated. Small for gestational age (SGA) was defined as birthweight < 10th centile for GA and sex; appropriate for gestational age (AGA) as birthweight ≥ 10th centile including large-for-gestational age (LGA) infants with birthweight ≥ 90th centile. Stunting and wasting were defined as length and weight/length ratio <3rd centile for GA and sex respectively. The anthropometric measures were compared to INTERGROWTH standards for weight, length and occipito-frontal circumference for boys and girls born at GAs between 24+^0^ and 42+^6^ weeks (20, 21), and weight/length ratio between 33+^0^ to 42+^6^ (22). For the weight/length ratio of babies born between 24+^0^ and 32+^6^ weeks, the z-scores and centiles were manually calculated using coefficients (means and standard deviations) obtained from the INTERGROWTH-21st project team.

The tFEF in days was calculated as the difference between the day the infant tolerated 120ml/kg/day and the day of birth. The time to regain birthweight was the difference between the day the baby regained birthweight and the day of birth. Time to final outcome, equivalent to the length of hospital stay, was computed from the date of discharge or death and the date of birth.

Clinical variables were summarised as proportions for categorical variables and as medians and interquartile ranges (IQR) for continuous/interval variables. Pearson’s correlation was performed to test correlation between continuous interval measures while Mann-Whitney U and Kruskal-Wallis tests were used to test for associations between tFEF and independent variables; the former for variables with 2 independent groups and the latter for variables with several independent groups. Multiple linear regression models were developed to control for confounding factors. A P value <0.05 was considered statistically significant and the effect sizes with confident intervals were calculated for independent predictors of tFEF.

### Ethics approval

Ethics approval for collection of anonymised patient data was granted by the LSTM Research and Ethics Committee and from the Institutional Ethics Committee for each NNU. The institutions and (Protocol Numbers) are as follows:

Liverpool School of Tropical Medicine (18–0210)

Jaramogi Oginga Odinga Teaching and Referral Hospital (ERC.IB/VOL.1/510)

University College Hospital, Ibadan (UI/EC/18/0446)

Massey Street Children’s Hospital (LSHSC/2222/VOL.VI^B^/185),

Ahmadu Bello University Teaching Hospital (ABUTH/HZ/HREC/D37/2018)

Maitama District Hospital (FHREC/2018/01/108/19-09-18)

Lagos University Teaching Hospital Health Research Ethics Committee (AMD/DCST/HREC/APP/2514)

Kenya Medical Research Institute-Scientific and Ethics Review Unit (KEMRI/SERU/CGMR-C/120/3740)

## Results

Of the 2280 babies enrolled in the NeoNuNet database, 1172 (51.4%) were preterm and 484 (21.2%) were both preterm and VLBW.

### Neonatal Characteristics

Of preterm/VLBW infants, 226 (46.7%) were male. The median (IQR) birthweight was 1.2 (1.1, 1.3) kg while the median GA was 30 (28, 32) weeks. A third of the preterm VLBW infants (117/393; 29.7%) were SGA, 42% of whom were disproportionately small (wasted) and about a fifth (73/393; 18.6%) were stunted; 53 and 85 infants were missing their birthweight and/or length at birth respectively. Classification by GA, birthweight and appropriateness of growth for GA and sex are presented in S1 Table. Twenty-six (6%) LGA infants were included with the AGA group because of their small numbers. Fourteen (3%) babies had congenital abnormalities (S2 Table).

### Feeding Practices

One hundred and thirty-six (28.0%) infants died before feeds could be commenced. Amongst the remaining 348 infants, median time to first feed was 46 (IQR: 27, 72) hours and only 29 (8.3%) infants were fed within the first hour of life. Mothers’ own milk was the most common type of first feed (270, 77.6%) then preterm formula (68, 19.5%); standard formula was rarely used (5, 1.4%). During admission, mother’s own milk remained the dominant type of feed; 203 infants were exclusively fed mothers’ own milk giving an exclusive breastfeeding rate of 58.3% but there was considerable mixed feeding (113; 32.5% infants). Twenty-four (6.9%) and 2 (0.6%) infants used preterm formula and standard formula exclusively respectively throughout the course of their admission.

Two hundred and forty-nine (71.6%) infants were initially tube fed via the orogastric/nasogastric route while 199 (57.2%) and 159 (45.7%) were fed by cup/cup-and-spoon and direct breastfeeding respectively.

Of the 348 infants who commenced feeds, 263 (75.6%) tolerated full enteral feeds (120ml/kg/day). Overall, full enteral feeds were established by a median time of 8 (4.5, 12) days but tFEF varied markedly between NNUs ranging between 1 – 32 days (Fig 1). Infants regained their birthweight by a median of 12 (7, 16) days while the median length of hospital stay among survivors was 27 (20, 38.5) days. There was a moderate positive correlation between tFEF and time to regain birthweight (r = .357, p < .001; Fig 2), and length of hospital stay (r = .303, p < .001; Fig 3). Time to regain birthweight also has a moderate, though greater, correlation with length of hospital stay (r = .378, p < .001; Fig 4).

**Figure 1:**
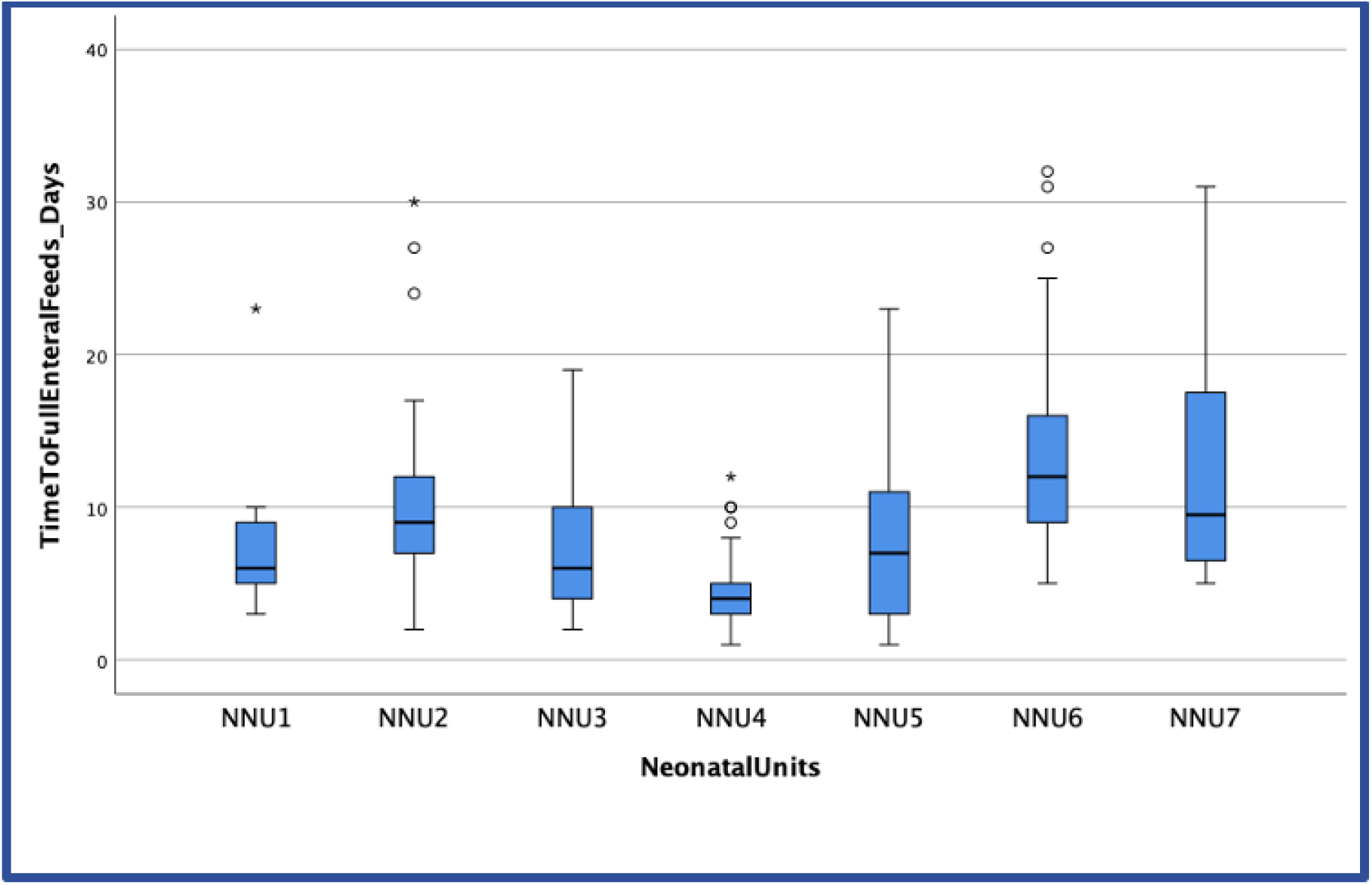
Boxplot of time to full enteral feeds by NNU. The horizontal line represents the median tFEF; the box represents the interquartile range; the whiskers represents the first and last quartiles tFEF. ∘ represent outlier and * extreme outliers

**Figure 2:**
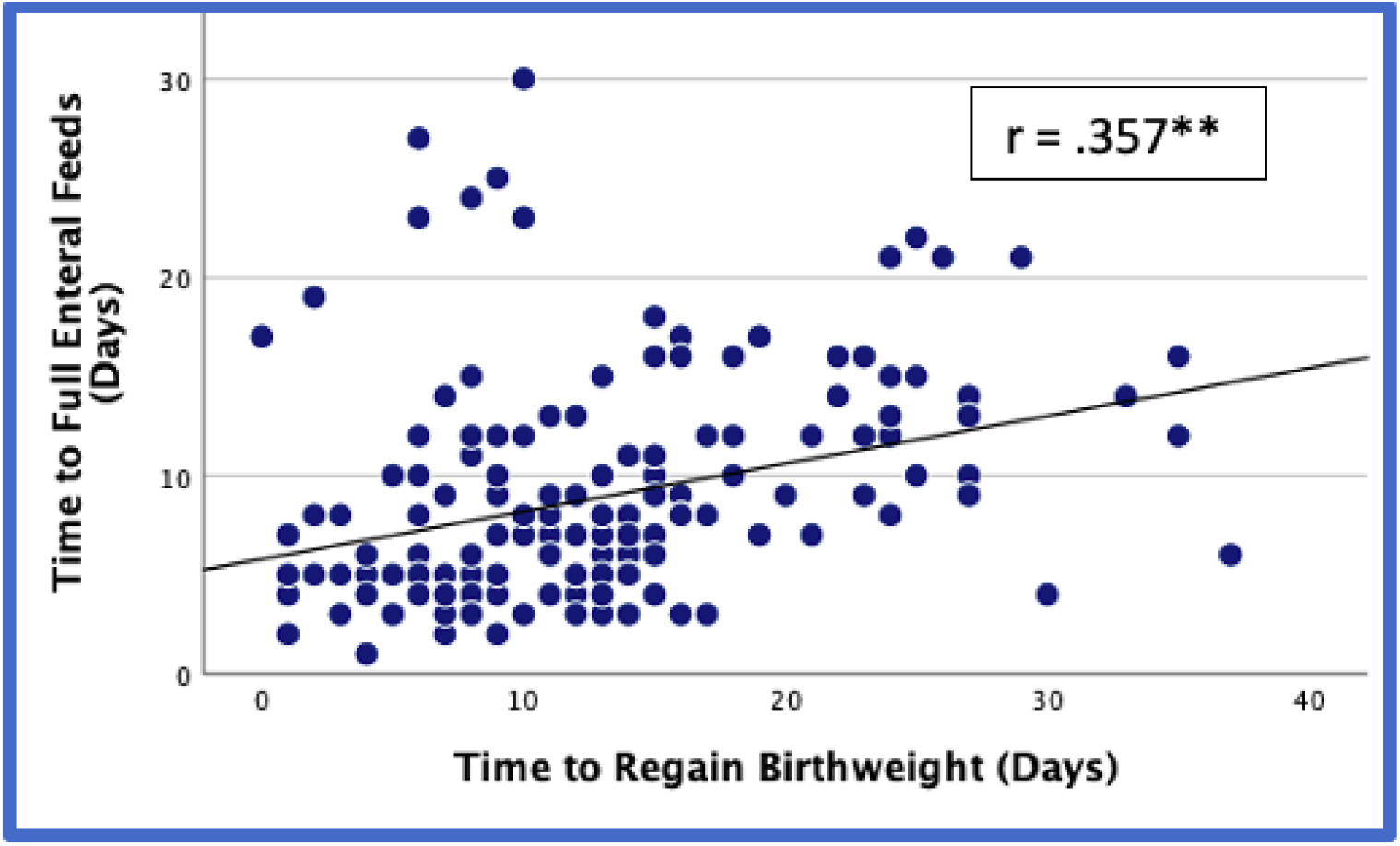
Correlation between tFEF and time to regain birthweight. **Correlation significant at the P≤0.01

**Figure 3:**
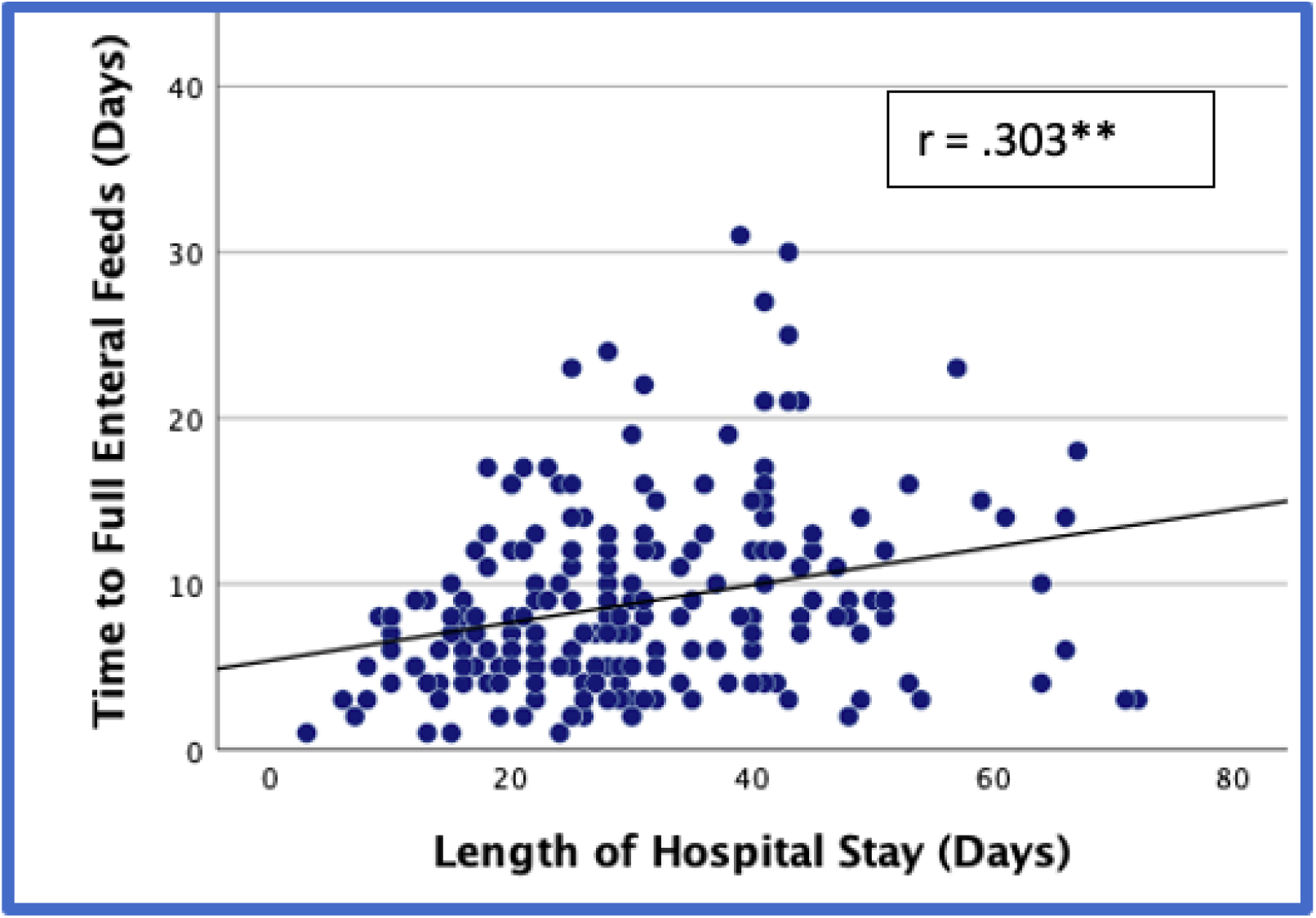
Correlation between tFEF and length of hospital stay. **Correlation significant at the P≤0.01

**Figure 4:**
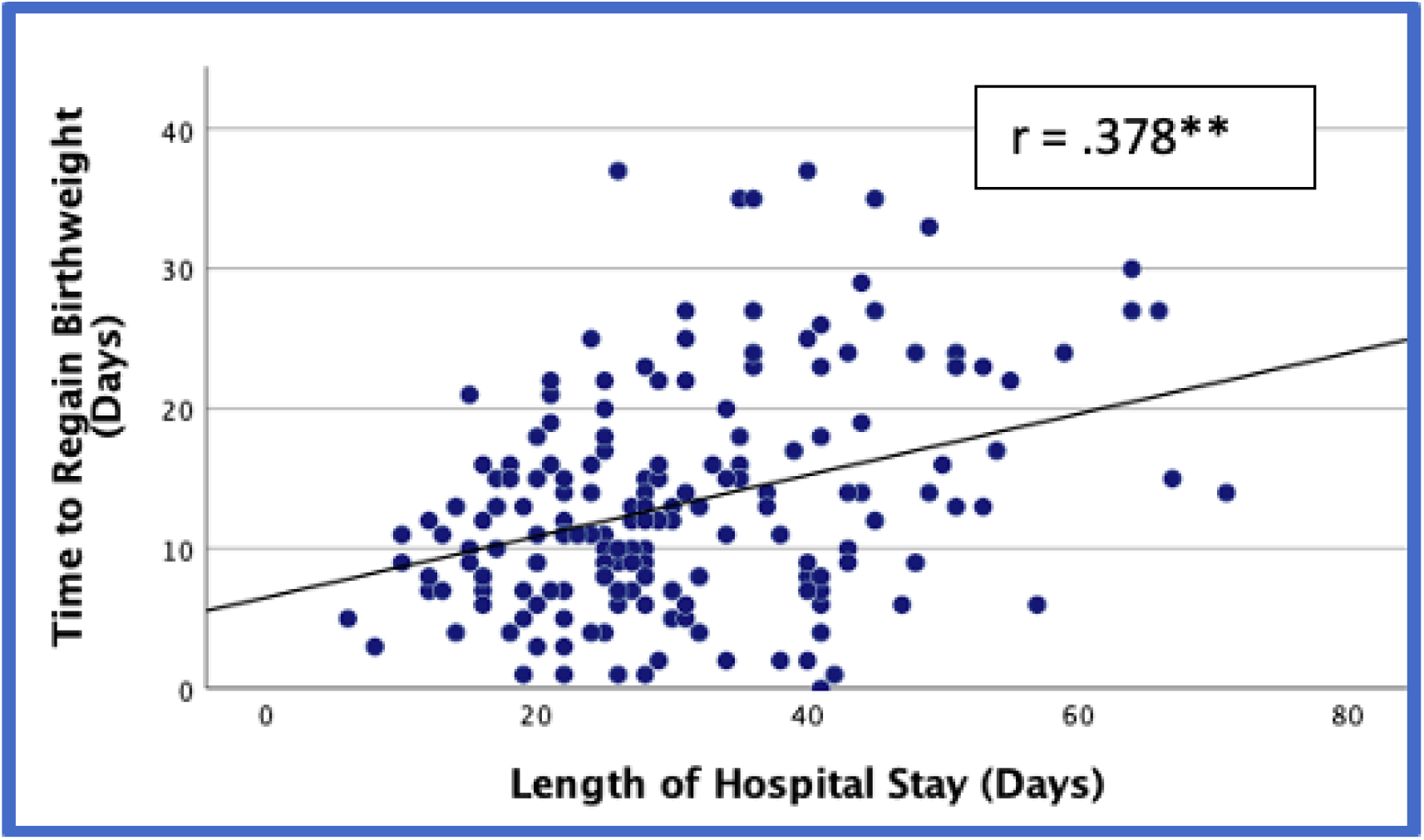
Correlation between time to regain brithweight and length of hospital stay. **Correlation significant at the P≤0.01

Overall, only 92 (19%) infants from 4 NNUs in Nigeria received parenteral amino acid infusions; one NNU used them routinely with 74% of their preterm and VLBW infants receiving parenteral amino acids for an average of 7 days. Parenteral lipids were only used on one patient in Nigeria.

### Morbidity and Mortality

Forty (8%) infants had perinatal asphyxia, 255 (53%) had varying degrees of respiratory distress, 217 (45%) developed sepsis and 22 (5%) developed abdominal conditions including feed intolerance and NEC. All the patients who had respiratory distress received oxygen therapy via modified bCPAP (118, 46%), free flow through nasal prongs (102, 40%) or high flow (37, 14%); oxygen therapy lasted for a median (IQR) of 3 (2, 7.75) days. The overall mortality rate was 46% (222/484) with half of the deaths occurring by 2 days and three-quarters by 6 days (S1 Fig). Case fatality rates were 62.5%, 49.0%, 46.5% and 59.0% for asphyxia, respiratory distress, sepsis and abdominal conditions respectively.

### Association between clinical variables and time to full enteral feeds

In univariate analysis, starting feeds earlier was associated with a shorter time to establish FEF (P<0.001). In pairwise comparisons with adjusted p-values, there was a significant reduction in tFEF when feeds were commenced on the 1st day of life compared to the 3rd (P =0.000, r = -.57), 4th (P = 0.000, r = -.48), 5th (P =.013, r = - .38) or 6th (P =0.004, r = -.42) days. There was no significant difference in tFEF between starting feeds on the 1st compared to the 2nd day of life (P =.660, r = -.23; Fig 5). Data were not collected on the volume of initial feed or the rate of advancement. There was no association between type of feed, sex or anthropometric indices and tFEF. Amongst common morbidities, the occurrence of an abdominal condition was associated with a longer tFEF (P=0.004). Conversely, respiratory distress was associated with a statistically significant shorter tFEF (P=0.001) (Table 1).

**Table 1:**
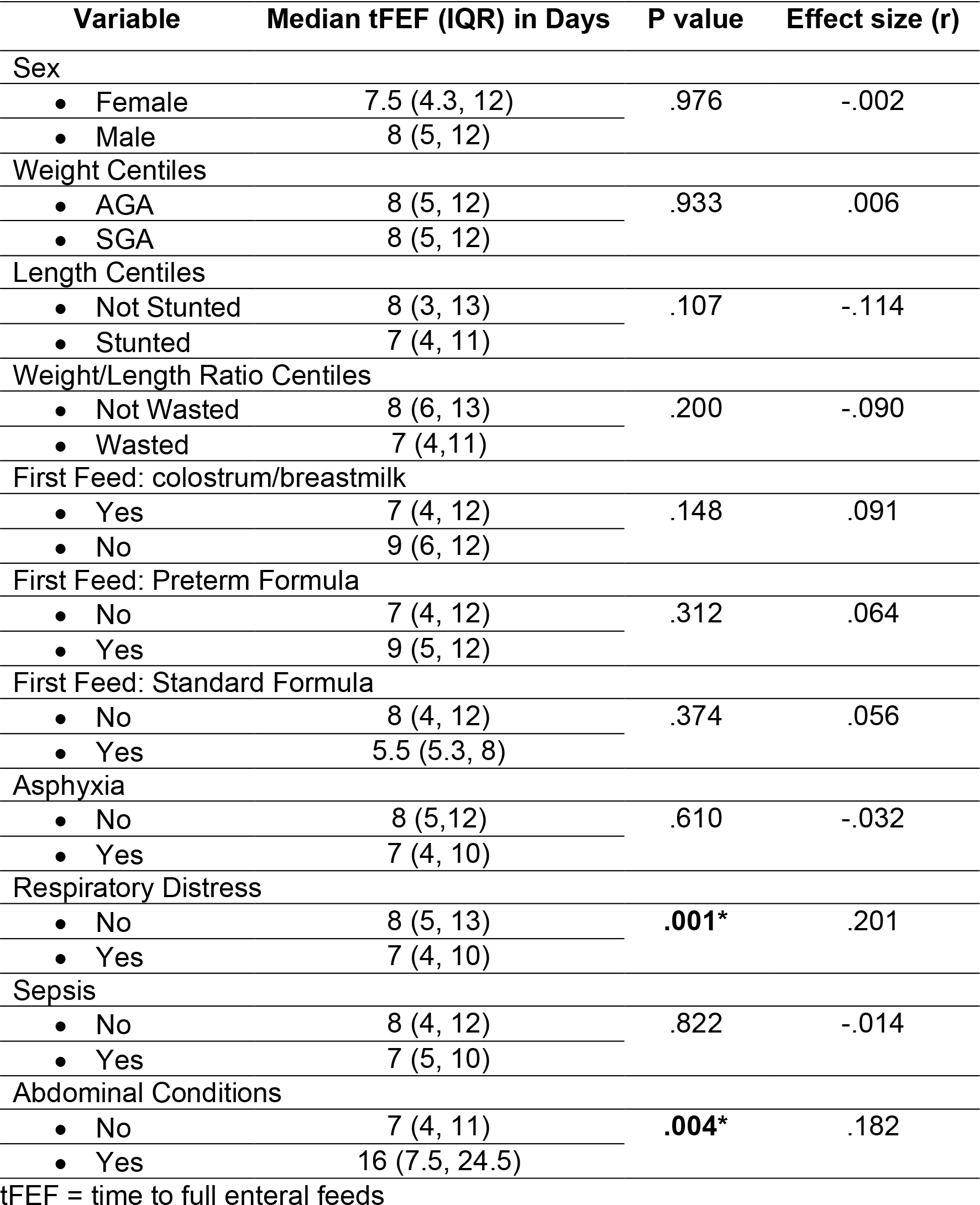
Associations between demographic and clinical variables and time to full enteral feeds; Univariate Analysis.

| Variable | Median tFEF (IQR) in Days | P value | Effect size (r) |
| --- | --- | --- | --- |
| Sex |  |  |  |
| • Female | 7.5 (4.3, 12) | .976 | -.002 |
| • Male | 8 (5, 12) |  |  |
| Weight Centiles |  |  |  |
| • AGA | 8 (5, 12) | .933 | .006 |
| • SGA | 8 (5, 12) |  |  |
| Length Centiles |  |  |  |
| • Not Stunted | 8 (3, 13) | .107 | -.114 |
| • Stunted | 7 (4, 11) |  |  |
| Weight/Length Ratio Centiles |  |  |  |
| • Not Wasted | 8 (6, 13) | .200 | -.090 |
| • Wasted | 7 (4,11) |  |  |
| First Feed: colostrum/breastmilk |  |  |  |
| • Yes | 7 (4, 12) | .148 | .091 |
| • No | 9 (6, 12) |  |  |
| First Feed: Preterm Formula |  |  |  |
| • No | 7 (4, 12) | .312 | .064 |
| • Yes | 9 (5, 12) |  |  |
| First Feed: Standard Formula |  |  |  |
| • No | 8 (4, 12) | .374 | .056 |
| • Yes | 5.5 (5.3, 8) |  |  |
| Asphyxia |  |  |  |
| • No | 8 (5,12) | .610 | -.032 |
| • Yes | 7 (4, 10) |  |  |
| Respiratory Distress |  |  |  |
| • No | 8 (5, 13) | **.001\*** | .201 |
| • Yes | 7 (4, 10) |  |  |
| Sepsis |  |  |  |
| • No | 8 (4, 12) | .822 | -.014 |
| • Yes | 7 (5, 10) |  |  |
| Abdominal Conditions |  |  |  |
| • No | 7 (4, 11) | **.004\*** | .182 |
| • Yes | 16 (7.5, 24.5) |  |  |

tFEF = time to full enteral feeds

**Figure 5:**
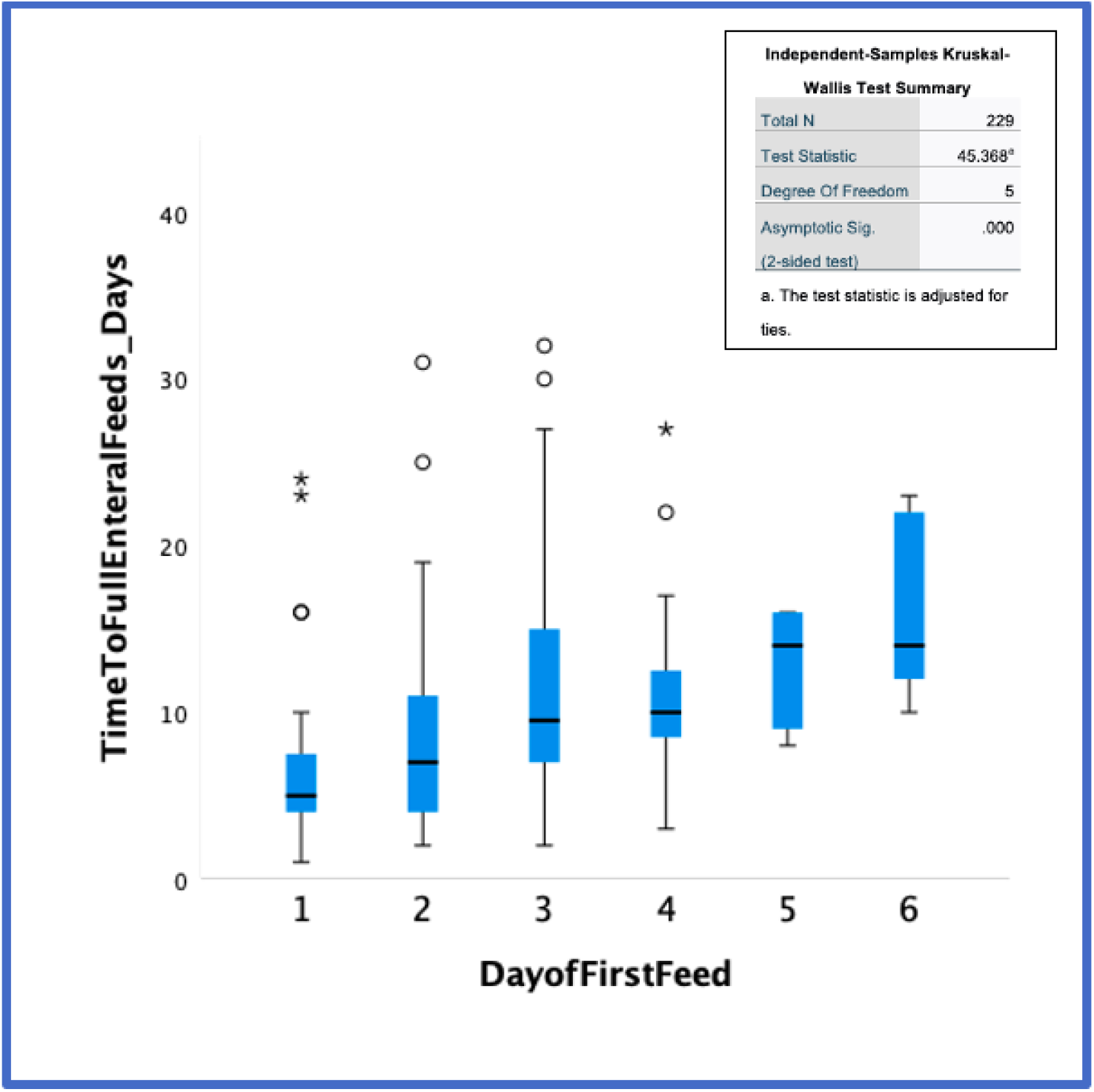
Median Time to Full Enteral Feeds according to Day of First Feed. The horizontal lines represents the median tFEF; the box represents the interquartile range of tFEF; the lower and upper whiskers represents the first and last quartiles tFEF respectively. º represent outlier tFEF and * extreme tFEF

Variables that were found to be independent predictors of tFEF on multiple linear regression analysis were day of first feed (unstandardised B 1.75; 95%CI 1.16, 2.34; P<0.001), respiratory distress −1.89 (−3.27, −0.51; P=.007) and NEC 4.54 (1.16, 7.92; P=0.009) (Table 2). This meant that for every 24-hour delay in commencing feeds (after the first 48hours), tFEF increased by 1.78 (95%CI; 1.09, 2.49) days; every day on oxygen therapy for respiratory distress decreased tFEF by 3.06 (−4.94, −1.18) days and for every day of being managed for NEC, tFEF was delayed by 4.34 (0.37, 8.34) days.

**Table 2:** Associations between demographic and clinical variables and time to full enteral feeds; Multiple Linear Regression analysis.

| Predictor Variable | Unstandardised Coefficient B<br>(95% CI) | P Value |
| --- | --- | --- |
| Constant | 5.71<br>(3.99, 7.43) | <.001 |
| Day of first feed | 1.75<br>(1.16, 2.34) | <.001 |
| Respiratory distress | -1.89<br>(-3.27, -0.51) | .007 |
| Abdominal<br>Conditions | 4.54<br>(1.16, 7.92) | .009 |

## Discussion

In this analysis, tFEF was significantly associated with time of first feed and the occurrence of respiratory distress and abdominal conditions in multiple regression analysis.

The NeoNuNet defined full enteral feeds as 120ml/kg/day, but other studies have used 120 and 150 ml/kg/day (11), 140 ml/kg/day (23) and 150-160ml/kg/day (24). The overall median time to full enteral feeds in this study was 8 (IQR: 4.5,12) days, De Waard et al reported that tFEF ranged from 8-33 days for full enteral feeds of 120ml/kg/day (11); although their analysis included data from 13 NNUs, only one was from Africa (Nigeria); median tFEF was 11 days for the sole Nigerian hospital. In the same study, median tFEF was 26 days (for 120ml/kg/day) in 5 NNUs in South China that used infant formula predominantly, feeds were commenced between the 2^nd^ to 5^th^ day of life and feed advancement was conservative.

Time to full enteral feeds in this study varied significantly across participating NNUs. This variation is unlikely due to differences in case mix as the analysis was restricted to preterm and VLBW infants and the frequency of common morbidities was similar across the NNUs. Differences between NNUs in feeding protocols likely contributed to variations in tFEF. Only centres in Kenya had written feeding guidelines and these advised slow feed advancement rates of 10 - 20ml/kg/day (13). This emphasises the need for standard protocols to reduce variation in practices to improve care. Adherence to an agreed protocol would also facilitate comparisons between units.

This analysis confirms the finding of other studies that the later feed is commenced, the later tFEF is attained (11). Feeds were commenced on the second or third day of life across the participating NNUs. Enteral feeds may be delayed because mothers’ own milk is not available, as a unit practice or due to the clinical status of the newborn. This analysis did not find any difference in tFEF among infants who were commenced on mothers’ own milk, preterm formula or standard formula. Kreissl et al, working in Vienna, comparing prospective observational data with retrospective medical records, found a decrease of 4 days in tFEF when feeds were commenced with single donor human milk compared to preterm formula; both groups commenced feeds on the first day life (23).

Clinical conditions that could delay the commencement of feeds include the need for bag-and-mask ventilation, perinatal asphyxia or respiratory distress. Perinatal asphyxia did not have a significant effect on tFEF in this analysis but interestingly, infants who had respiratory distress attained FEF significantly earlier than those who did not. This may be because these infants were placed on oxygen therapy (free flow or via modified bubble continuous positive airway pressure; bCPAP) and adequate oxygenation prevents hypoxia and the dive reflex. The dive reflex results in abnormal intestinal oxygenation, which is a risk factor for NEC (7, 8). Better oxygenation for this cohort of preterm infants with respiratory distress may have improved tolerance of feeds and hastened the attainment of full enteral feeds.

Although 30% of the preterm VLBW babies were SGA, this was not associated with tFEF. Although infants who were wasted (asymmetric SGA) and those who were stunted tended to have shorter tFEF the difference did not reach significance.

It would appear that feeds were delayed as a result of the feeding practice in use in the participating NNUs (13) – a practice informed by the fear of the preterm VLBW developing NEC (7, 8, 10). However, there is strong evidence from systematic reviews that delay in commencement, and slow advancement of enteral feeds do not decrease the incidence of NEC. Rather, data suggest these practices may result in up to 2 more cases of NEC, 3 more deaths and 15 more cases of feed intolerance, and it may also result in 5 to 6 more cases of sepsis per 100 VLBW infants (15-19). Slow advancement also results in increased hospital stay of up to 6 days (18, 19) which increases the direct and indirect cost of care particularly in LMICs where hospital fees are borne out-of-pocket (25). Sub-group analyses based on the type of feed (breastmilk vs. formula vs. mixed feeds) or if the infants were ELBW (extreme low birth weight; <1kg) and/or SGA did not show a significant association with the incidence of NEC.

Predictably, infants who had feed intolerance and/or NEC attained full enteral feeds as much as 8 days later than those who did not. The clinical signs and management of feed intolerance and NEC necessitate the interruption and/or discontinuation of feeds in addition to the use of antibiotic therapy and surgical intervention (7) which would delay the achievement of full enteral feeds as was found in this analysis. The association of an abdominal condition with delay in attaining tFEF was significant despite the low incidence of this complication in this cohort. The low NEC rates may be as a result of a third of the cohort dying before they could be commenced on enteral feeds (a key risk factor in the pathogenesis of NEC). The de Waard study did not find a significant difference in NEC rates across NICUs in South China and Europe/Oceania/North America/Africa despite differences in feeding protocols, but it did not report on any statistical tests of association between tFEF and NEC or other short-term outcomes (11).

## Strengths

One of the strengths of this study is that it involved prospective data collection which allowed for a more objective and complete dataset. Another strength is the large number of preterm and VLBW infants recruited from NNUs providing different levels of care, in resource-limited settings in sub-Saharan Africa which may make the results more generalisable. The routine clinical data collected provides an opportunity to identify gaps in clinical care.

## Limitations

There were a number of limitations with this analysis. Firstly, the details on initial feed volume, the rate of advancement, whether tube feeds were given in boluses or continuously or the frequency of bolus feeds were not collected, and these may have also impacted on tFEF. However, these data were collected in a related survey that was conducted among 37 Neonatologists/Paediatricians working in Nigeria and 13 in Kenya (13). The survey found that feeds were initiated with and advanced at 10-20ml/kg/day and were given in 2-3 hourly boluses by over 70%, 80% and 100% of respondents respectively.

In addition, variations in feeding practices between NNUs could not be accounted for, making it difficult to evaluate the true effects of birth anthropometry, GA and morbidity on tFEF. A further study with standardised feeding protocols across these NNU’s would provide greater certainty.

## Conclusion

Our findings among hospitalised preterm/VLBW infants admitted to NNUs in African settings strongly suggest that delays in initiating enteral feeds is associated with delays in achieving full enteral feeds resulting in longer hospital stay. The implementation of evidence-based, standardised feeding protocols is needed urgently with further research to evaluate their impact.

## Data Availability

All relevant data are within the manuscript and its Supporting Information files

## Acknowledgements

We would like to thank our colleagues who contributed to the clinical care and collection of data at all the neonatal units in the Network including at the University College Hospital, Ibadan; Lagos University Teaching Hospital; Massey Street Children’s Hospital, Lagos; Ahmadu Bello University Teaching Hospital, Zaria and Maitama District Hospital, Abuja in Nigeria. In Kenya they include the Jaramogi Oginga Odinga Teaching and Referral Hospital, Kisumu and The Kilifi County Referral Hospital. We would also like to thank our colleagues at the Nigerian Society of Neonatal Medicine, the Kenya Paediatric Association and the Ministries of Health in Nigeria and Kenya, who provided us with support and advice as we were setting up this study. We thank the mothers for participating in this study with their infants.

## Neonatal Nutrition Network members

Isa Abdulkadir (Ahmadu Bello University, Zaria, Nigeria); Ismaela Abubakar (Liverpool School of Tropical Medicine, Liverpool, UK); Abimbola E Akindolire (College of Medicine, University of Ibadan, Nigeria); Olusegun Akinyinka (College of Medicine, University of Ibadan, Nigeria); Stephen J Allen (Liverpool School of Tropical Medicine, Liverpool, UK); Pauline EA Andang’o (Maseno University, Kenya); Graham Devereux (Liverpool School of Tropical Medicine, Liverpool, UK); Chinyere Ezeaka (Lagos University Teaching Hospital, Nigeria); Beatrice N Ezenwa (Lagos University Teaching Hospital, Nigeria); Iretiola B Fajolu (Lagos University Teaching Hospital, Nigeria); Zainab O Imam (Lagos State University Teaching Hospital, Lagos, Nigeria); Kevin Mortimer (Liverpool School of Tropical Medicine, Liverpool, UK); Martha K Mwangome (KEMRI Wellcome Trust Research Programme, Kilifi, Kenya); Helen M Nabwera (Liverpool School of Tropical Medicine, Liverpool, UK); Grace M Nalwa (Jaramogi Oginga Odinga Teaching and Referral Hospital, Kisumu, Kenya & Maseno University, Kenya); Walter Otieno (Jaramogi Oginga Odinga Teaching and Referral Hospital, Kisumu, Kenya & Maseno University, Kenya); Macrine Olwala (Jaramogi Oginga Odinga Teaching and Referral Hospital, Kisumu, Kenya); Alison W Talbert (KEMRI Wellcome Trust Research Programme, Kilifi, Kenya); Nicholas D Embleton (Newcastle University, Newcastle, UK); Olukemi O Tongo (College of Medicine, University of Ibadan, Nigeria); Dominic D Umoru (Maitama District Hospital, Abuja, Nigeria); Janneke van de Wijgert (University of Liverpool, Liverpool, UK); Melissa Gladstone (University of Liverpool, Liverpool, UK).

## Open access

This is an open access article distributed in accordance with the Creative Commons Attribution 4.0 Unported (CC BY 4.0) license, which permits others to copy, redistribute, remix, transform and build upon this work for any purpose, provided the original work is properly cited, a link to the licence is given, and indication of whether changes were made. See: https://creativecommons.org/ licenses/by/4.0/.

## Supporting information

**S1 Table. Characteristics of Preterm and VLBW Infants**

**S2 Table. Congenital Abnormalities**

**S1 Fig. Time of death of infants**

